# Clinical management, epidemiology, and recurrence of human cystic echinococcosis in a secondary care level hospital in an endemic area of the Andes in Sicuani, Cusco, Peru

**DOI:** 10.1101/2024.09.12.24313559

**Authors:** Roberto Pineda-Reyes, Miguel M. Cabada, Maria L. Morales, Jan Hattendorf, Paola Vergaray, Ruben Bascope, Jakob Zinsstag

## Abstract

Peru has the highest incidence of human cystic echinococcosis (CE) in South America and most cases are reported in the Central and Southern Peruvian Andes, including the Cusco region. We reviewed medical records of patients with CE admitted between 2010-2019 to a level-2 hospital in Sicuani, which serves the Canchis province of Cusco. We collected data on surgical management and disease recurrence. Records from 115 patients were included, their median age was 25 years (IQR, 16-46) and 60% were female. Most patients (68.7%) presented with a single liver cyst. The Gharbi classification was used in 75/107 (70.1%) patients with an ultrasound and in 91/142 cysts (64.8%). Thirty-one (34.1%) were classified as Gharbi I, 33 (36.3%) Gharbi II, 21 (23.1%) Gharbi III, and 6 (6.6%) Gharbi IV. Pre-surgical complications were reported in 41.7%. One hundred two patients underwent surgery. In 39.2% a secondary CE prevention measure was documented, 5.9% had two, and one had the three recommended measures. Post-surgical cyst recurrence was reported in 16.5% at a median 32.3 months (IQR, 3.9-46.6) and readmissions in 12.7%. In the multivariable analysis, having ≥3 cysts (OR 6.26, 95% CI 1.10-39.85) was associated with post-surgical cyst recurrence. Advanced and complicated CE and post-surgical recurrence were common in Sicuani. Standardization of ultrasound staging-guided clinical management along with secondary CE prevention measures could effectively decrease the disease burden linked to clinical care, improving outcomes and decreasing costs. Ultimately, only integrated control at the source of infection including hand hygiene, secure disposal of offal, avoidance of dog feeding offal, and deworming of dogs can reduce transmission and lead to elimination.

## Introduction

Cystic echinococcosis (CE) is a zoonosis caused by the metacestode stage of the dog tapeworm *Echinococcus granulosus* (1)*. E. granulosus* has a global distribution, being endemic in South America, North and East Africa, Central Asia and China (2). Peru has the highest incidence of CE in South America, accounting for 84.8% of all reported cases in year 2018 (3). The district of Sicuani in the Cusco region is a highly endemic area in the Southern Peruvian Andes. Up to 45% of CE cases referred to tertiary care hospitals in Cusco are from Sicuani (4). A population survey using focus ultrasound to detect abdominal CE in four communities of the Cusco region found that 78% of patients diagnosed with CE were from Sicuani (5). The economic and urban expansion of the Sicuani city have created areas where unregulated livestock slaughter activities, stray dogs, and humans interact closely. Prompt diagnosis and appropriate treatment of cases in Sicuani are crucial to address the burden of CE in the healthcare system.

The WHO-Informal Working Group on Echinococcosis (WHO-IWGE) ultrasound staging system is used to diagnose and guide the clinical management of liver CE (6). However, many low- and middle-income countries (LMIC) have not implemented the proposed WHO ultrasound staging which may expose patients to unnecessary procedures (7). In developing countries, CE has a high post-surgical recurrence risk, with rates that range between 4.7% and 16.4% (8–11). Clinical characteristics such as the cyst size, number of cysts, and presence of complications have been associated with disease severity and recurrence (10, 12, 13). Treatment precautions to avoid cyst rupture and intraabdominal seeding may decrease the risk of recurrence related to surgical care.

The recommendations to avoid cyst rupture and secondary CE include the use of surgical field protection with hypertonic saline-soaked drapes, injection of a scolicidal agent into the cyst, and prescription of pre-surgical albendazole (ABZ) (6). Increasing evidence shows that appropriate standardization of surgical procedures may be associated with lower risk of recurrence (14–16). A prospective study found no mortality and zero recurrence at 24 months after standardized partial cystectomy, which was in part attributed to careful avoidance of intraabdominal spillage and prescription of perioperative ABZ.(16) In addition, the “watch-and-wait” approach is recommended as the standard care for inactive cysts to spare asymptomatic patients from unnecessary treatment (17, 18).

We conducted a review of medical records of patients admitted during a 10-year period to a low-complexity hospital in the city of Sicuani in Cusco, Peru, to describe the CE presentation, management, and post-surgical recurrence in a highly endemic area.

## Methods

### Study design and setting

This is a review of medical records from patients hospitalized at the Alfredo Callo Rodriguez (ACR) Hospital in Sicuani. This hospital is a level-2 public healthcare center that provides specialty medical care to patients from the Canchis and Canas provinces of Cusco (19). During March 2024, we collected data from all patients admitted to the hospital for CE between January 1, 2010, and December 31, 2019. The pre-COVID-19 period was selected to avoid collecting information that could reflect gaps in care or practice of CE treatment in Sicuani because of the pandemic public health emergency response. This timeframe also coincides with the renovation and expansion of surgical services in the hospital in 2010.

The city of Sicuani is the capital of the Canchis province in the Cusco region at an elevation of 3,550 meters above sea level. The Sicuani district has an urban population of 47,400 and a rural population of 10,400 habitants (20). Sicuani is a commerce center for farmers of the surrounding areas and is the second largest city in the Cusco region.

### Study procedures

Medical record numbers of patients with CE were identified using ICD-10 codes (B67.0-B67.9) assigned as the discharge diagnosis through the hospital’s statistics department. Surgical logbooks were handsearched to ensure all surgical CE cases were retrieved. Demographic and clinical data were collected via ODK software (© 2022 Get ODK Inc., California, USA) using an electronic case report form, which was piloted at the start of data collection to ensure data accuracy.

### Definitions

*Pre-surgical complications* were defined as documented evidence of cyst-related infectious processes, fistulas, mass effect, allergic manifestations, hemorrhage, or trauma before surgical treatment. One patient who presented to the hospital with abdominal pain and a 13-week pregnancy was also considered complicated before surgery. The three assessed *secondary CE prevention measures* included surgical field protection with drapes soaked in hypertonic saline, injection of hypertonic saline into the cyst prior to evacuation, and the prescription of pre-surgical ABZ (6). Hemoglobin values were adjusted for age, sex, pregnancy status, and district of residence elevation in order to define *mild, moderate* or *severe anemia* (21). *Post-surgical recurrence* was defined as the development of a new cyst at or near the site of the previously removed cyst and/or within the cavity (e.g., peritoneum, pleura) where cyst(s) were operated on. The *time-to-recurrence* was estimated as the time from the surgery date until the date of recurrence diagnosis. In one case the date of recurrence was documented only as “2024” and it was assigned “January-1-2024” as the corresponding diagnosis date. *New cyst* was defined as a newly diagnosed cyst in a location different from other operated cysts. *Readmission* was defined as return to inpatient care related to CE after discharge. *Readmission rate* was calculated by dividing the number patients with at least one readmission by the total number of hospitalization discharges.

### Annual cumulative incidence calculation

The annual cumulative incidence of new hospital admissions was calculated dividing the number of new CE hospitalizations per year by the total population at risk each year. The population at risk was defined as the total population of the Canas and Canchis provinces per year. Population data for years 2015 to 2019 was extracted from a repository database of the Peruvian Ministry of Health and data for years 2009 to 2014 was extracted from a third party vendor database, CPI Peru (22, 23).

### Statistical analysis

Data analysis and visualization were performed using the R software, version 4.3.2 (R Foundation for Statistical Computing, Vienna, Austria). Means with standard deviation (SD), medians with interquartile range (IQR), and frequencies were used to assess the distributions of demographic, epidemiological, and clinical characteristics of patients. A multivariable logistic regression analysis was performed to determine independent associations with the outcome variable “post-surgical recurrence”. We pre-selected the variables “age”, “sex”, and added further variables to the multivariable model based on p<0.15 on univariable analyses with the outcome variable. A P value of <0.05 was considered statistically significant. Confidence intervals were calculated at the 95% level.

### Ethical Statement

This study was reviewed and approved by the Ethics Committee of Northern and Central Switzerland (Ethikkommission Nordwest-und Zentralschweiz, EKNZ, No. AO_2023-00096) and the Institutional Ethics Committee (Comite Institucional de Etica en Investigacion, No. 213622) of the Universidad Peruana Cayetano Heredia in Lima, Peru.

## Results

### Demographic and clinical characteristics

One hundred and twenty-five medical record numbers were identified. Six were duplicated, 2 were misrecorded as CE, and 2 had missing charts. In total, 115 unique medical records from patients with CE were included in the study (**Table 1**). The median age was 25 years (IQR, 16-46) and 60% were female. Most patients (103, 89.6%) were residents of the Canchis province and 73.9% lived in the Sicuani district. Fifty-six patients (48.7%) were referred from primary healthcare centers and 57.1% of these were referred from the Pampaphalla health center.

**Table 1.**
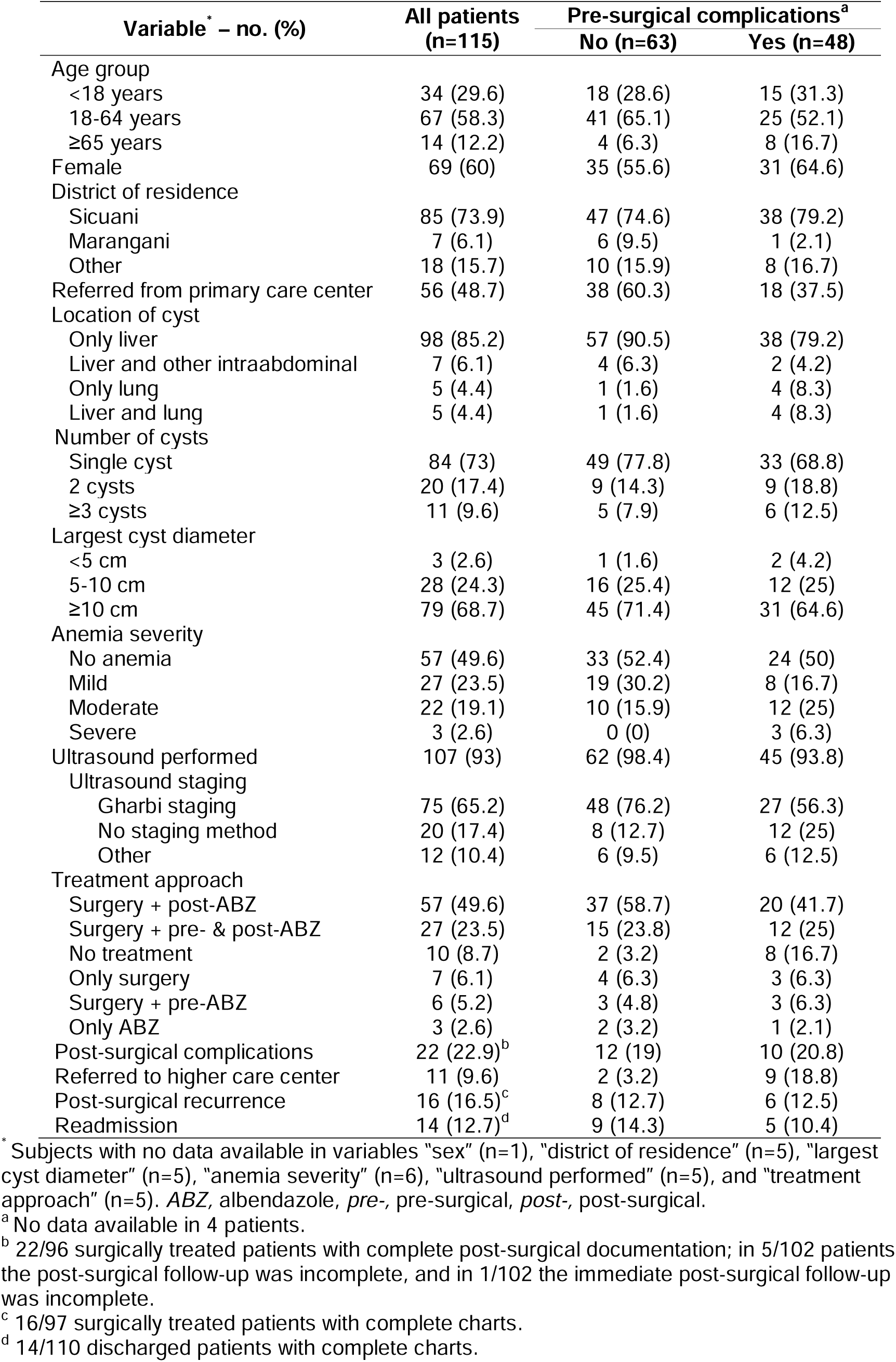
General characteristics of 115 patients hospitalized for cystic echinococcosis between 2010 and 2019 at the Alfredo Callo Rodriguez Hospital of Sicuani, Cusco, Peru.

One hundred eighty-five echinococcal cysts were diagnosed in the 115 patients, 169 were diagnosed upon presentation to the hospital and 16 during readmissions. These involved the liver, lungs, stomach, spleen, duodenum, gallbladder, diaphragm, adnexa, bladder, and colon. Most (142, 84%) were located in the liver, 16 within the abdominal cavity, and 11 in the lungs. Four patients had disseminated liver and intraabdominal cysts. The maximum number of lung cysts per patient was 2. The median diameter of the largest liver cyst per patient was 12.8 cm (IQR, 9.2-15.0) and for lung cysts was 8.5 cm (IQR, 4.5-12.6). Seventy-seven (67%) patients had a liver cyst of ≥10 cm in diameter.

Most patients (98, 85.2%) presented with cysts only in the liver, 7 (6.1%) with both liver and intraabdominal cysts, 5 (4.3%) with liver and lung cysts, and 5 (4.3%) with only lung cysts. Most patients presented with a single liver cyst (79, 68.7%). Forty-eight (41.7%) cases presented with complicated cysts before surgery, 31 (27%) had one pre-surgical complication, and 17 (14.8%) had two or more. The most common pre-surgical complications were infection of the cyst content (17, 14.9%), cysto-biliary fistulas (9, 7.8%), liver abscess (9, 7.8%), cyst rupture (8, 7%), and peritonitis (5, 4.3%) (**Table 2**). A summary of the clinical presentation characteristics and laboratory results is shown in **Supplementary Table 1S**.

**Table 2.** Pre-surgical, intraoperative, and post-surgical complications by anatomical location of cysts.

| Complication type* | n (%) |
| --- | --- |
| **Pre-surgical (n=48, 41.7%<sup>a</sup>)** |  |
| Infectious (n=29, 25.2%) |  |
| Purulent cyst fluid | 17 (14.8) |
| Abscess | 9 (7.8) |
| Peritonitis | 5 (4.4) |
| Pneumonia | 5 (4.4) |
| Cholecystitis | 2 (1.7) |
| Cholangitis | 1 (0.9) |
| Mechanical (n=23, 20%) |  |
| Fistula | 11 (9.6) |
| Cysto-biliary | 9 (7.8) |
| Cysto-pleural-bronchial | 1 (0.9) |
| Cholecysto-gastric | 1 (0.9) |
| Cyst rupture or vomica | 10 (8.7) |
| Mass effect<sup>b</sup> | 5 (4.4) |
| Respiratory (n=5, 4.4%) |  |
| Respiratory insufficiency | 5 (4.4) |
| Hemorrhagic (n=3, 2.6%) |  |
| Hemoptysis | 2 (1.7) |
| Hemorrhagic cyst fluid | 1 (0.9) |
| Allergic (n=3, 2.6%) |  |
| Rash/urticaria | 3 (2.6) |
| Other (n=3, 2.6%) |  |
| Pregnancy | 1 (0.9) |
| Abdominal trauma | 1 (0.9) |
| **Intraoperative (n=2, 2%<sup>c</sup>)** |  |
| Anaphylactic shock | 1 (1.0) |
| Cardiac arrest | 1 (1.0) |
| Hemoperitoneum | 1 (1.0) |
| Spleen laceration | 1 (1.0) |
| **Post-surgical (n=22, 22.9%<sup>d</sup>)** |  |
| Residual collection | 14 (14.6) |
| Abscess | 3 (3.1) |
| Biliary leakage | 3 (3.1) |
| Cutaneous fistula | 2 (2.1) |
| Adhesions | 1 (1.0) |
| Hemorrhage | 1 (1.0) |
| Post-cardiac arrest | 1 (1.0) |
| Sepsis | 1 (1.0) |
| Wound infection | 1 (1.0) |

\* Patients may have more than one complication.
<sup>a</sup> 48/115 total patients.
<sup>b</sup> Biliary obstruction (n=3), bowel obstruction (n=1), abdominal compartment syndrome (n=1).
<sup>c</sup> 2/102 surgically treated patients.
<sup>d</sup> 22/96 surgically treated patients with complete post-operative documentation.

### Ultrasound diagnosis

One hundred seven patients (93%) had a diagnostic ultrasound. In 75 (70.1%), a liver ultrasound was performed and used for staging according to the Gharbi classification (6), and in 20 (18.7%) a liver ultrasound was performed but no staging was reported. Twelve patients had lung cysts, disseminated disease, complicated cysts, or unclear or missing ultrasound reports precluding the use of a cyst diagnostic and management classification. In total, 91 (64.8%) of the 142 liver cysts were staged using the Gharbi classification. Thirty-one (34.1%) were Gharbi I, 33 (36.3%) Gharbi II, 21 (23.1%) Gharbi III, and 6 (6.6%) GharbiIV. Seven (6.1%) patients were diagnosed with inactive cysts, including one who had a Gharbi IV cyst diagnosed intraoperatively.

### Treatment, secondary CE prevention measures, and post-surgical complications

Only patients with liver and intraabdominal cysts underwent surgery in Sicuani (102, 88.7%). Ninety-five of the 102 patients (93.1%) underwent open surgery, 6 (5.9%) laparoscopic surgery, and one laparoscopic surgery converted to open laparotomy. The most common surgical techniques were cystectomy pl us drainage (74, 72.5%) and pericystectomy plus drainage (9, 8.8%) (**Supplementary Table 2S**). In one case a Gharbi IV cyst was found and not excised and two patients with inactive cysts (2%) underwent excision. In one case the liver cyst was not found during the surgery. None of the patients had a percutaneous procedure performed. In 18 (17.6%) patients, the surgical report did not describe the location of the cyst in the liver. Seventy (68.6%) patients had right liver lobe cysts, 26 (25.5%) left liver lobe cysts, and 12 (11.8%) had cysts in both liver lobes. In 46 (45.1%) patients the liver segment VIII was involved, in 32 (31.4%) the segment VII, in 24 (23.5%) the VI, in 17 (16.7%) the IV, in 12 (11.8%) the V, in 10 (9.8%) the II, in 8 (7.8%) the III, and in one patient the segment I.

Forty (39.2%) patients had one secondary CE prevention measure documented, 6 (5.9%) had two, and only one had the three recommended measures. Pre-surgical ABZ was used in 33 (32.4%) cases, injection of hypertonic saline before cyst opening was documented in 16 (15.7%) cases, and surgical field protection with hypertonic saline soaked drapes was used in 6 (5.9%) cases.

ABZ was the only antiparasitic prescribed. In 57 (55.9%) patients, ABZ was prescribed after surgery, in 27 (26.5%) before and after surgery, in 7 (6.9%) no ABZ was prescribed, in 6 (5.9%) ABZ was prescribed only before surgery, and in 5 cases the timing of the prescription was unknown. The median post-surgical duration of ABZ treatment prescribed was 30 days (IQR, 22-34) and the median pre-surgical duration of ABZ treatment prescribed was 2 days (IQR, 1-4).

Post-surgical complications occurred in 22/96 (22.9%) patients with complete post-operative documentation. The most common were the development of residual fluid collections (14, 14.6%), post-surgical abscess (3, 3.1%), and biliary leakage (3, 3.1%) (**Table 2**). The three patients with post-surgical liver abscess were readmitted to the hospital.

### Post-surgical recurrence, readmission, and annual cumulative incidence

One patient required a second surgical intervention within the same hospitalization for post-surgical biliary leak, and one required intensive care due to post-surgical hemodynamic instability. Eleven (9.6%) patients were referred to a higher level of care, 9 for treatment of pulmonary CE, 1 for post-surgical shock, and 1 for potential need of post-surgical intensive care. Of these, 10 patients were transferred to the Hospital Regional del Cusco and 1 patient to the Hospital Antonio Lorena in Cusco. Post-surgical recurrence was documented in 16/97 (16.5%) patients. The median time to recurrence after surgery was estimated at 32.3 months (IQR, 3.9-46.6). The multivariable logistic regression analysis showed that having ≥3 cysts compared to a single cyst (50% vs 15.1%, OR=6.26, 95% CI 1.10-39.85) was associated with post-surgical recurrence (**Figure 1 and Supplementary Table 3S**).

**Figure 1.**
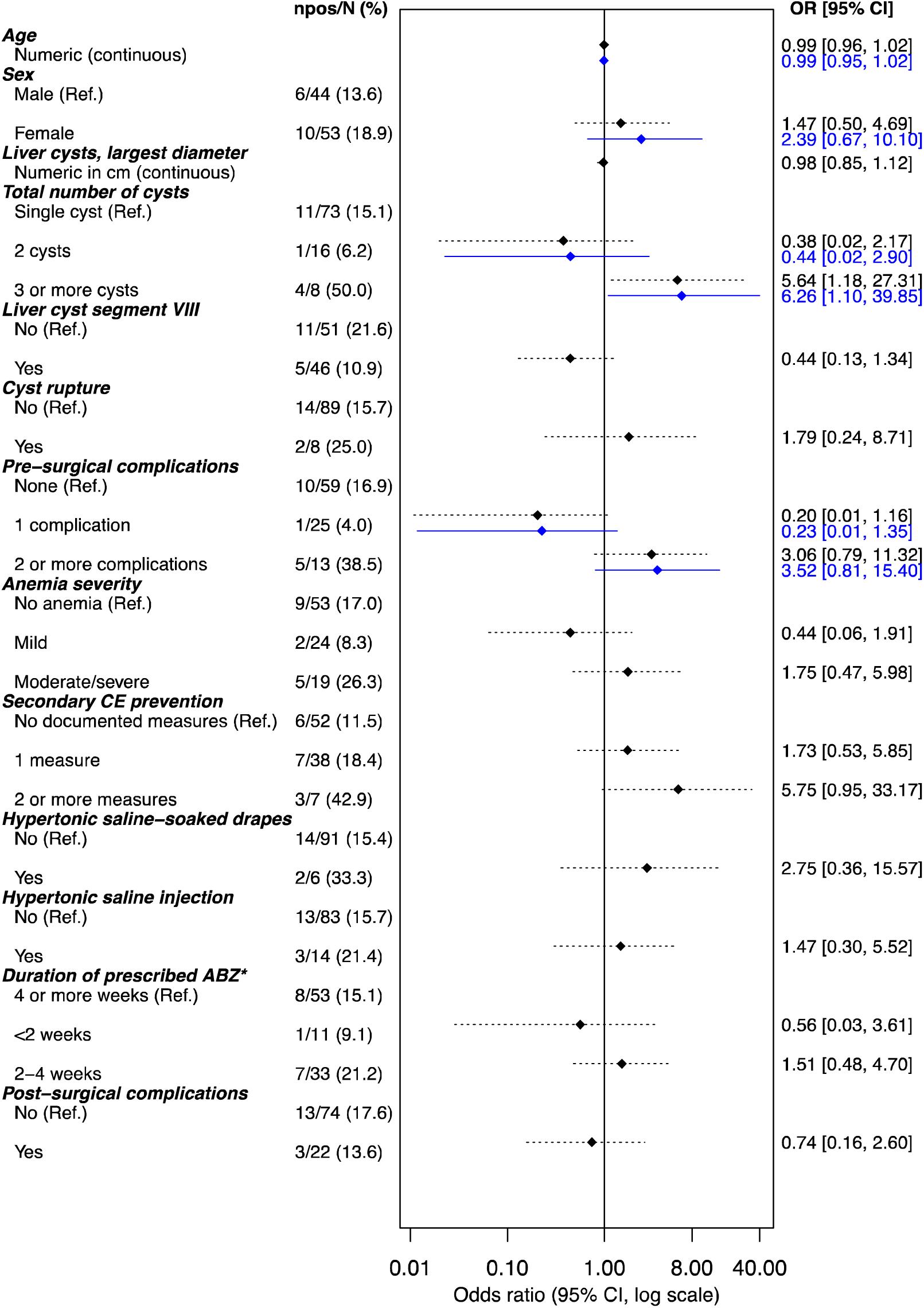
Forest plot showing the univariable and multivariable logistic regression analyses of variables associated with post-surgical recurrence. Odds ratios and confidence intervals in black represent univariable logistic regression models for each independent variable. Odds ratios and confidence intervals in blue represent the multivariable analysis. ABZ, albendazole, CI, confidence interval, *npos/N*, number with recurrence/total per category, OR, odds ratio, *Ref.,* reference category. *Including pre- and post-surgical treatment.

Fourteen (14/110, 12.7%) patients were readmitted to the hospital at least once, 6 did for cyst recurrence, 2 for treatment complications, 2 for complications of a previous cyst plus cyst recurrence, 3 for elective two-stage treatment, and one for a new cyst. One patient had 2 readmissions. Twelve out of the 14 patients with readmissions had active echinococcal cysts. The median age was 40 years (IQR, 26.5-62), 7 (58.3%) were female, and 5 (41.7%) were referred from the Pampaphalla health center. Single liver cyst was the presentation in 9/12 (75%) of patients with readmissions. Readmissions contributed with 16 additional cysts to the overall count for the 10-year study period. The annual cumulative incidence of new hospital admissions for CE at the ACR Hospital of Sicuani are reported in **Table 3**.

**Table 3.** Annual cumulative incidence of new hospital admissions for CE at the Alfredo Callo Rodriguez Hospital of Sicuani during the period 2010-2019.

| Year | Hospital admissions for new CE diagnosis |  |  | All hospital admissions due to CE* |  |  | Population at risk<sup>§</sup> | Cumulative incidence<br>×100,000 |
| --- | --- | --- | --- | --- | --- | --- | --- | --- |
|  | Patients | Echinococcal cysts |  | Admissions | Echinococcal cysts |  |  |  |
|  | n=115 (%) | n=169 (%) | Mean number of cysts per patient (SD) | n=126 (%) | n=185 (%) | Mean number of cysts per patient (SD) |  |  |
| 2019 | 21 (18.3) | 33 (19.5) | 1.57 (1.12) | 22 (17.5) | 35 (18.9) | 1.59 (1.10) | 144,607 | 14.5 |
| 2018 | 16 (13.9) | 21 (12.4) | 1.31 (0.79) | 17 (13.5) | 22 (11.9) | 1.29 (0.77) | 143,826 | 11.1 |
| 2017 | 16 (13.9) | 27 (16.0) | 1.69 (1.78) | 19 (15.1) | 30 (16.2) | 1.58 (1.64) | 143,059 | 11.2 |
| 2016 | 12 (10.4) | 17 (10.1) | 1.42 (1.17) | 15 (11.9) | 20 (10.8) | 1.33 (1.05) | 142,266 | 8.4 |
| 2015 | 20 (17.4) | 31 (18.3) | 1.55 (0.83) | 23 (18.2) | 38 (20.5) | 1.65 (1.07) | 141,444 | 14.1 |
| 2014 | 9 (7.8) | 11 (6.5) | 1.22 (0.44) | 9 (7.1) | 11 (6.0) | 1.22 (0.44) | 141,900 | 6.3 |
| 2013 | 5 (4.4) | 9 (5.3) | 1.8 (0.84) | 5 (4.0) | 9 (4.9) | 1.8 (0.84) | 142,300 | 3.5 |
| 2012 | 9 (7.8) | 11 (6.5) | 1.22 (0.44) | 9 (7.1) | 11 (6.0) | 1.22 (0.44) | 142,800 | 6.3 |
| 2011 | 5 (4.4) | 7 (4.1) | 1.4 (0.89) | 5 (4.0) | 7 (3.8) | 1.4 (0.89) | 143,200 | 3.5 |
| 2010 | 2 (1.7) | 2 (1.2) | 1.0 (0.00) | 2 (1.6) | 2 (1.1) | 1.0 (0.00) | 143,600 | 1.4 |

\*Including readmissions, but not counting one patient readmitted in 2020 for a liver cyst, one patient readmitted in 2020 for multiple liver and intraabdominal cysts, and two patients readmitted for drainage of post-surgical liver abscesses.
<sup>§</sup>Combining the Canas and Canchis provinces total population.
CE, cystic echinococcosis, SD, standard deviation.

## Discussion

Our study describes the clinical characteristics and management of CE at a low-complexity hospital in an endemic area of the Southern Peruvian Andes. Most CE patients requiring admission to this hospital were part of the economically active population and presented often with advanced disease (cysts ≥10 cm in diameter) and/or pre-surgical complications. Despite the common use of ultrasound for the evaluation of CE, the WHO-IWGE system was not used for the diagnosis and management of liver CE in any of the cases. Instead, the Gharbi classification was used on a limited number of cysts. Secondary CE prevention measures were documented in less than half of the surgical procedures. One in six patients had recurrence of CE after surgery and one in eight patients were readmitted to the hospital at least once. Our data suggest that educational interventions could improve the medical and surgical management of CE at the ACR Hospital of Sicuani, thus decreasing the morbidity associated with disease recurrence and surgical procedures.

Between 2016-2018, the Peruvian Ministry of Health reported a CE cumulative incidence of 29.2-45 cases per 100,000 in the Cusco region, which is one of the highest in the country (24). During the same period, our cumulative incidence calculation ranged between 8.4-11.2 new admissions to the ACR Hospital per 100,000. These numbers are also higher than most other regions in Peru, even though they do not include outpatient consultations, cases diagnosed in primary healthcare centers, and cases from the Social Security Health System (EsSalud) or private healthcare centers. The ACR Hospital of Sicuani serves the populations of the Canas and Canchis provinces. These provinces have large young age populations, which are affected by CE. The period between infection and clinical presentation, or ‘latent period’, typically lasts many years in CE. This suggests that active transmission occurs in Sicuani and that it happens early in life. A large case series of hospitalized patients in two regions of Spain showed a distribution skewed towards the older age groups with only 2% cases in patients younger than 19 years (25, 26). The authors attributed this age distribution in part to the successful interruption of transmission by control programs based on stray dogs elimination and safe disposal of livestock offal (27). A similar scenario was described in Chile, where most reported cases between 2018-2022 were diagnosed among patients between 45-74 years (28). Efforts to control CE transmission in the Canas and Canchis provinces have been limited to the treatment of dogs in rural areas and sporadic screening of children. A concerted effort with a One Health approach is crucial to formulate effective control strategies in the area.

In our study, most patients had liver cysts and 41.7% presented to the hospital with complications. This is a high proportion of complications, similar to the 40.3% reported in a study in Salamanca, Spain (29). Compared to the latter, our study had slightly lower percentage of mechanical complications (20% vs 25%); however, we encountered a higher proportion of infectious complications (25.2% vs 17.8%), cyst rupture (8.7% vs 2.9%), and advanced disease (cysts ≥10 cm in 68.7% versus 45%) (29). These discrepancies could be due to a selection bias related to the hospital level of complexity, our lower proportion of lung cysts, or due to differences in healthcare access. Another retrospective series in the Junin region in the Central Peruvian Andes also reported over 50% cysts ≥10 cm, but their proportion of pre-surgical complications was significantly lower (4%) likely due to including cases from outpatient services and private hospitals (30).

We could not establish a significant association between pre-surgical complications and CE recurrence. Other studies with larger samples were also unable to demonstrate associations between infectious, mechanical, or allergic complications and CE recurrence (9, 29). This could indicate that the risk of recurrence depends on other clinical variables associated with surgical or post-surgical care (9). Large cyst diameter has been proposed as a marker of severity and risk factor for recurrence (12). Despite the large cyst diameters found in our patients compared to other studies, an association between this variable and pre-surgical complications, recurrence, or readmission was not detected (11, 13, 29). We did observe an association between higher number of cysts and risk of post-surgical recurrence. Number of cysts has been identified as risk factor for complications and recurrence (10, 13). Pacifico and colleagues found that patients with ≥3 cysts had a four-fold higher risk for pre-surgical complications in a secondary case-control analysis of a study in the Central Andes of Peru (13). Similarly, having ≥3 cysts was an independent risk factor for recurrence in a retrospective study of 672 surgically treated patients in Morocco (10). A large parasite burden may lead to subclinical cyst rupture and seeding in some patients before hospital admission or during surgical interventions.

Ultrasound staging was documented in two-thirds of those with liver cysts. Ultrasound CE diagnosis and staging is not yet implemented in many LMIC (7). The WHO-IWGE staging system was not used for the management of CE in our study sample. This classification serves as a clinical management guide to consider medical and invasive (percutaneous, surgical) interventions, or a ‘watch and wait’ approach (6). Patients with liver cysts who underwent surgery presented with Gharbi I or II in our study, and some could have been candidates for percutaneous treatment (31). However, this treatment option is not available in endemic areas of Peru. Patients with Gharbi III cysts in our study had a greater proportion of recurrence compared to the other stages. Surgery is the treatment of choice for Gharbi III cysts due to high recurrence rates reported in percutaneous treatment studies (31, 32). Multiple daughter cysts within a solid matrix, as seen in Gharbi III, may be more difficult to access and drain likely missing smaller cysts which may not get sufficient scolicidal drug exposure within the matrix, increasing the risk of treatment failure (32, 33). Early ultrasound screening along with implementation of WHO-IWGE staging and percutaneous procedures may decrease costs, unnecessary procedures, and hospitalization time in Sicuani.

We found a higher post-surgical recurrence rate in comparison to other studies from different global regions (8, 10, 11, 34–37). A minority of patients had documentation of recommended interventions to prevent spillage of cyst contents during surgery. In some cases, surgical field drapes were soaked on normal saline or iodine rather than hypertonic saline. In others, the instillation of a scolicidal agent into the cyst was performed after opening and evacuation. An association between spillage-prevention measures and post-surgical recurrence could not be determined. In addition, patients with documented adequate spillage-prevention measures had a higher proportion of recurrence. This could be explained by differences in documentation or care in patients with more complicated disease. In these patients, the surgeons may have performed a more detailed documentation of interventions or used spillage prevention more often. Although careful implementation of these measures has been associated with no recurrence at 24 months in prospective studies (16), it is unclear whether other measures used at the ACR Hospital are effective. We could not find an association between recurrence and the timing of ABZ prescription (e.g., pre-surgical, post-surgical, or both) or the duration of ABZ treatment but our power to find these association was probably limited by our sample size. In addition, there was no documentation of medication adherence, and some patients might have had problems to afford or take antiparasitics consistently. Pre-surgical ABZ is a core measure for secondary prevention in liver CE (31). Prospective studies assessing the timing and dosing of antiparasitics as well as the appropriateness of spillage-prevention measures during surgery may help close this information gap.

Our study had limitations that should be acknowledged. First, this is a retrospective review of medical charts with a small sample size. Missing information in certain variables such as ultrasound staging did not allow testing their association to the outcome. Second, as the hospital had a digitalized database of medical record numbers only since 2017, cases between 2010-2016 were manually searched in the surgical logbooks. Therefore, patients hospitalized during this period who did not undergo surgery are missing in our cohort. Third, the description of symptoms, complications, and management of patients in our study represent the clinical judgement and documentation of different surgeons in Sicuani during a long time span. These may not concur with the standard of care in other contexts and generalizations should be carefully assessed. Additionally, surgical reports documentation differed among surgeons, which precluded an analysis on the association of surgical techniques and recurrence. Finally, the lack of follow-up in some patients may have caused an underestimation of cyst recurrence.

In conclusion, advanced and complicated liver CE and post-surgical recurrence were common among hospitalized patients at the ACR Hospital in the city of Sicuani in Cusco, Peru. Practical measures including standardization of ultrasound-guided clinical management, secondary CE prevention measures, and follow-up could decrease the CE burden linked to clinical care, improving outcomes and decreasing costs. Ultimately, the contextual ecological cycle of transmission must be established in Sicuani, as only an integrated control at the source of infection involving interventions such as hand hygiene, secure disposal of offal, avoidance of dog feeding offal, and deworming of dogs can reduce transmission and lead to elimination. This requires a One Health approach and a socio-ecological strategy analysis (38), similar to rabies control (39).

## Data Availability

All data produced in the present study are available upon reasonable request to the authors.

## Acknowledgements

We thank the Alfredo Callo Rodriguez Hospital’s Administration Office as well as the personnel of the Statistics and Archives Department for their assistance during the study period.

## Authors’ details

**RP-R:** Swiss Tropical and Public Health Institute, associated to University of Basel, Allschwil, Switzerland, Kreuzstrasse 2, 4123 Allschwil, Switzerland; **MMC:** Division of Infectious Diseases, University of Texas Medical Branch, Galveston Texas, 301 University Boulevard, Rte. 0435, Galveston, TX 77555-0435, USA, and Cusco Branch – Alexander von Humboldt Tropical Medicine Institute, Universidad Peruana Cayetano Heredia, Jr. Jose Carlos Mariategui J-6, Wanchaq 08002, Cusco, Peru; **MLM:** Cusco Branch – Alexander von Humboldt Tropical Medicine Institute, Universidad Peruana Cayetano Heredia, Jr. Jose Carlos Mariategui J6, Wanchaq 08002, Cusco, Peru; **JH:** Swiss Tropical and Public Health Institute, associated to University of Basel, Allschwil, Switzerland, Kreuzstrasse 2, 4123 Allschwil, Switzerland; **PV:** Departamento de Cirugia, Hospital Alfredo Callo Rodriguez – Sicuani, Av. Manuel Callo Zevallos Nro. 519, Sicuani, Cusco, Peru; **RB:** Programa de Control de Enfermedades Zoonoticas, Gerencia Regional de Salud de Cusco, Prolongacion Avenida de la Cultura 1525, Cusco 08003, Peru; **JZ:** Swiss Tropical and Public Health Institute, associated to University of Basel, Allschwil, Switzerland, Kreuzstrasse 2, 4123 Allschwil, Switzerland.

