## Supplementary_Table_1S_2S_3S for "Clinical management, epidemiology, and recurrence of human cystic echinococcosis in a secondary care level hospital in an endemic area of the Andes in Sicuani, Cusco, Peru"

**Supplementary Table 1S.** Clinical presentation characteristics by anatomical location of cysts.

| **Variable^*^** | **Only liver (n=98)** | **Liver and other intraabdominal (n=7)** | **Liver and lung (n=5)** | **Only lung (n=5)** |
| --- | --- | --- | --- | --- |
| Median age (IQR) | 28 (17-47.5) | 21 (13-41) | 18 (6-80) | 18 (8-23) |
| Female – no. (%) | 57 (58.2) | 3 (42.9) | 5 (100) | 4 (80) |
| Symptoms – no. (%) |  |  |  |  |
| Abdominal pain | 94 (95.9) | 6 (85.7) | 2 (40) | 0 (0) |
| Nausea | 26 (26.5) | 2 (28.6) | 0 (0) | 0 (0) |
| Vomiting | 22 (22.4) | 1 (14.3) | 0 (0) | 0 (0) |
| Abdominal mass | 6 (6.1) | 0 (0) | 0 (0) | 0 (0) |
| Fever | 5 (5.1) | 0 (0) | 1 (20) | 3 (60) |
| Shortness of breath | 2 (2) | 0 (0) | 3 (60) | 4 (80) |
| Cough | 0 (0) | 0 (0) | 4 (80) | 5 (100) |
| Hemoptysis | 0 (0) | 0 (0) | 1 (20) | 1 (20) |
| Vomica | 0 (0) | 0 (0) | 1 (20) | 1 (20) |
| Median duration of symptoms (IQR) | 31 (8-92) | 19.5 (14.3-120.8) | 13 (7-15) | 20 (8-62) |
| Number of cysts – no. (%) |  |  |  |  |
| Single cyst | 79 (80.6) | 0 (0) | 0 (0.0) | 5 (100) |
| 2 cysts | 16 (16.3) | 1 (14.3) | 3 (60.0) | 0 (0) |
| ≥3 cysts | 3 (3.1) | 6 (85.7) | 2 (40.0) | 0 (0) |
| Median largest cyst diameter (IQR) | 13 (9.6-15.1) | 9.5 (7.4-13.1) | 10.9 (10.7-12) | 5 (4-9.8) |
| Median unadjusted hemoglobin (g/dl) (IQR) | 15 (13.5-16.3) | 15.1 (14.7-15.5) | 13 (12.6-13.5) | 13.6 (12-13.6) |
| Anemia severity – no. (%) |  |  |  |  |
| No anemia | 49 (50.0) | 5 (71.4) | 1 (20) | 2 (40) |
| Mild | 23 (23.5) | 1 (14.3) | 2 (40) | 1 (20) |
| Moderate | 19 (19.4) | 0 (0) | 2 (40) | 1 (20) |
| Severe | 2 (2) | 0 (0) | 0 (0) | 1 (20) |
| Median WBC count (×10^9^/L) (IQR) | 9.2 (7.5-12.3) | 10.3 (8.5-14.4) | 11.5 (7.6-12.1) | 10.2 (8-11) |
| Median eosinophil count (cells/μL) (IQR) | 128 (0-300) | 352 (176-548) | 0 (0-470) | 35 (0-81.6) |
| Median platelet count (×10^9^/L) (IQR) | 315.2 (298.4-350.7) | 389 (340-392) | 337 (260-378) | 492 (357-550) |
| Median AST (units/L) (IQR) | 10.1 (7-18.8) | 9 (8-9.5) | 39 (22-40.5) | NA |
| Median ALT (units/L) (IQR) | 12 (9-17.7) | 11 (9.5-11) | 36 (22.5-46.5) | NA |
| Median total bilirubin (mg/dL) (IQR) | 1.02 (0.84-1.39) | 0.89 (0.86-1.02) | 0.24 (0.18-0.31) | NA |
| Post-surgical recurrence – no. (%) | 13 (13.3) | 3 (42.9) | 0 (0) | 0 (0) |
| Readmission – no. (%) | 12 (12.2) | 2 (28.6) | 0 (0) | 0 (0) |

^*^ Subjects with no data available in variables “sex” (n=1), “duration of symptoms” (n=12), “largest cyst diameter” (n=5), “pre-surgical complications” (n=4), “hemoglobin” (n=6), “anemia severity” (n=6), “WBC count” (n=7), “eosinophil count” (n=28), “platelet count” (n=18), “AST” (n=63), “ALT” (n=63), “total bilirubin” (n=59), “post-surgical recurrence” (n=5), and “readmission” (n=5).

*ALT,* alanine transaminase, *AST,* aspartate transaminase, *IQR,* interquartile range, *NA*, non-available, *WBC*, white blood cell.

**Supplementary Table 2S.** Surgical techniques performed among 102 patients during the study period. In one case the liver cyst could not be found, and the patient was discharged on albendazole.

| **Surgical procedure described** | **N=102 (%)** |
| --- | --- |
| Cystectomy plus drainage | 74 (72.5) |
| Cystectomy | 5 (4.9) |
| Partial cystectomy plus drainage | 2 (2) |
| Pericystectomy plus drainage | 9 (8.8) |
| Pericystectomy | 4 (3.9) |
| Laparoscopic cystectomy plus drainage | 3 (2.9) |
| Laparoscopic cystectomy | 1 (1) |
| Laparoscopic drainage of abscessed cyst | 1 (1) |
| Enucleation and cystectomy | 1 (1) |
| Cyst not excised | 1 (1) |
| Cyst not found | 1 (1) |

**Supplementary Table 3S.** Univariable logistic regression analysis of variables assessing their association with post-surgical recurrence.

| **Variable – no. (%)** | **Post-surgical recurrence^a^** | | **uOR** | **95% CI** | ***P* value** |
| --- | --- | --- | --- | --- | --- |
|  | **No (n=81)** | **Yes (n=16)** |  |  |  |
| Age, continuous^*^ | 24 (16-46) | 20 (16.8-32.5) | 0.99 | 0.96-1.02 | 0.60 |
| Sex |  |  |  |  |  |
| Male | 38 (86.4) | 6 (13.6) | Ref |  |  |
| Female | 43 (81.1) | 10 (18.9) | 1.47 | 0.5-4.69 | 0.49 |
| Liver cyst largest diameter in cm^*^ | 13 (9.2-15) | 11.7 (9.3-14.9) | 0.98 | 0.85-1.12 | 0.74 |
| Number of cysts |  |  |  |  |  |
| Single cyst | 62 (84.9) | 11 (15.1) | Ref |  |  |
| 2 cysts | 15 (93.7) | 1 (6.3) | 0.38 | 0.02-2.17 | 0.37 |
| ≥3 cysts | 4 (50) | 4 (50) | 5.64 | 1.18-27.31 | 0.03 |
| Involvement of liver segment VIII |  |  |  |  |  |
| Yes | 41 (89.1) | 5 (10.9) | 0.44 | 0.13-1.34 | 0.16 |
| No | 40 (78.4) | 11 (21.6) | Ref |  |  |
| Pre-surgical cyst rupture |  |  |  |  |  |
| Yes | 6 (75) | 2 (25) | 1.79 | 0.24-8.71 | 0.50 |
| No | 75 (84.3) | 14 (15.7) | Ref |  |  |
| Pre-surgical complications |  |  |  |  |  |
| No complications | 49 (83.1) | 10 (16.9) | Ref |  |  |
| 1 complication | 24 (96) | 1 (4) | 0.2 | 0.01-1.16 | 0.14 |
| ≥2 complications | 8 (61.5) | 5 (38.5) | 3.06 | 0.79-11.32 | 0.09^†^ |
| Anemia severity |  |  |  |  |  |
| No anemia | 44 (83) | 9 (17) | Ref |  |  |
| Mild | 22 (91.7) | 2 (8.3) | 0.44 | 0.06-1.91 | 0.33 |
| Moderate/severe | 14 (73.7) | 5 (26.3) | 1.75 | 0.47-5.98 | 0.38 |
| Secondary CE prevention measures |  |  |  |  |  |
| None documented | 46 (88.5) | 6 (11.5) | Ref |  |  |
| 1 measure | 31 (81.6) | 7 (18.4) | 1.73 | 0.53-5.85 | 0.36 |
| ≥2 measures | 4 (57.1) | 3 (42.9) | 5.75 | 0.95-33.17 | 0.05^¶^ |
| Hypertonic saline-soaked drapes |  |  |  |  |  |
| Yes | 4 (66.7) | 2 (33.3) | 2.75 | 0.36-15.57 | 0.27 |
| No | 77 (84.6) | 14 (15.4) | Ref |  |  |
| Hypertonic saline cyst injection |  |  |  |  |  |
| Yes | 11 (78.6) | 3 (21.4) | 1.47 | 0.3-5.52 | 0.59 |
| No | 70 (84.3) | 13 (15.7) | Ref |  |  |
| Prescription of ABZ |  |  |  |  |  |
| Only post-surgical | 50 (87.7) | 7 (12.3) |  |  |  |
| Pre- and post-surgical | 18 (66.7) | 9 (33.3) |  |  |  |
| None documented | 7 (100) | 0 (0) |  |  |  |
| Only pre-surgical | 6 (100) | 0 (0) |  |  |  |
| Duration of prescribed ABZ treatment |  |  |  |  |  |
| <2 weeks | 10 (90.9) | 1 (9.1) | 0.56 | 0.03-3.61 | 0.61 |
| 2-4 weeks | 26 (78.8) | 7 (21.2) | 1.51 | 0.48-4.70 | 0.47 |
| ≥4 weeks | 45 (84.9) | 8 (15.1) | Ref |  |  |
| Duration of prescribed ABZ in days (continuous) | 30 (21-34) | 28.5 (23.8-34) | 1.01 | 0.98-1.04 | 0.41 |
| Post-surgical complications, any |  |  |  |  |  |
| Yes | 19 (86.4) | 3 (13.6) | 0.74 | 0.16-2.6 | 0.67 |
| No | 61 (82.4) | 13 (17.6) | Ref |  |  |

^a^ N=97, including 102 surgically treated patients and excluding 5 patients with incomplete charts. *ABZ,* albendazole, *CE,* cystic echinococcosis, *uOR,* unadjusted odds ratio. ^*^Median (IQR).

^†^Overall model statistically significant (Likelihood Ratio Chi-square Test, *P*=0.024).

^¶^Overall model not statistically significant (Likelihood Ratio Chi-square Test, *P=*0.216).
